# Indian Interventional trials for COVID-19 drugs: Insights and Learnings

**DOI:** 10.1101/2021.05.16.21257299

**Authors:** Arati Ranade, Kirtee Wani, V. Premnath, Chitra Lele, Smita Kale

## Abstract

Since the COVID-19 pandemic began, India has substantially contributed to drug development and clinical research. Task Force on Repurposing of Drugs (TFORD) for COVID19 has tried to look at the overall position of India in terms of interventional clinical trials and highlight learnings which can prepare us to fight future pandemics in a better way. Trials registered on CTRI from March 2020 to December 2020 were considered for this purpose. From a total 409 trials registered, 108 focused on modern drugs. From 108 trials studied, 92 were randomized trials, 34 trials were sponsored by Indian Pharmaceutical industry, 23 were self-sponsored and 20 were sponsored by Research institutes and hospitals. Only 83 trials studied the repurposed drugs. An unfortunate revelation was that out of 108 trials, 79 showed as not yet recruiting. This highlights the urgent need for Government, Research institutions and Indian Pharmaceutical industries to break down silos and work together towards this common cause.

## Introduction

Coronavirus disease 2019 (COVID-19), a contagious disease caused by severe acute respiratory syndrome coronavirus 2 (SARS-CoV-2), was first identified in Wuhan, China, in December 2019. Since then, it has spread worldwide, leading the World Health Organization (WHO) to declare this as the public health emergency with an international concern and to term this outbreak a global pandemic^1^.

Many institutions and researchers worldwide are involved in finding pharmacological interventions to COVID-19 at the earliest. A remarkable achievement has been seen on this path through the development of vaccines in a short span of time. Nevertheless, considering the urgent requirement of precise therapeutics to fight this pandemic and the obvious limitations of vaccine use, the need for anti-viral agents, anti-inflammatory agents or monoclonal antibodies to fight this pandemic still exists. According to the data published by FDA’s Coronavirus Treatment Acceleration Program (CTAP), as on 31^st^ December 2020 there were 590+ drug development programs in planning stages, 400+ trials reviewed by FDA, 8 treatments currently authorised for emergency use and 1 treatment approved for COVID 19^2^.

Our literature survey reveals the availability of few studies capturing the position of COVID 19 clinical research in India and world-wide. A study by Vasantha Raju N., & Harinarayana N.S. attempts to provide a comprehensive overview on clinical trials that are being conducted in India on COVID-19^3^. Authors capture the data available on CTRI website till July 2020. They found “interventional studies were more compared to observational studies and that majority of the clinical trials were investigating AYUSH related interventions.

Another review by Sujit Bhattacharya *et*.*al*^4^ maps the research papers and clinical trials from 22 May 2020 to 2 June 2020 to provide an informed view of the status of the research and drug development activity.

An article by Jaykaran Charan *et*.*al*^5^ provides information regarding potential interventions for COVID 19 from India till July 2020.

A comprehensive overview of the COVID-19 research in India including design aspects, through the clinical trials registered in the Clinical Trials Registry -India (CTRI) till June 5, 2020 is elaborated in a manuscript by Maulik *et. al*^6^.

A review by Mark P. Lythgoe^7^ published in June 2020 captures registered interventional clinical trials for the treatment and prevention of COVID-19 to provide insight into the global response to this pandemic.

A review by Mehta *et. al*^8^ presents cross-sectional analysis of clinical trials available till 26 March 2020 for the treatment of COVID-19 that were registered in the USA or in countries contributing to the WHO’s International Clinical Trials Registry Platform.

To the best of our knowledge there are no manuscripts compiling the clinical trial data from India for the total period of March 2020 to December 2020. Our objective here is not only to provide the overall position of India in terms of interventional clinical trials related to modern drugs on COVID 19 but also to highlight learnings which can prepare us to fight future pandemics in a better way.

Since the pandemic began, India has substantially contributed to drug development and clinical research. Principal Scientific Adviser to the Government of India Dr K Vijay Raghavan constituted the Task Force for Repurposing of Drugs (TFORD) for COVID-19, under the aegis of Science and Technology core group. This Task Force was constituted to track the latest and continuously evolving information on drug candidates, carry out an inter-disciplinary and inter-institutional assessment of drug candidates, anticipate and strengthen a translation ecosystem for drug development and to support informed decision making. The Venture Center on the CSIR-NCL campus serves as the information nerve center for this Task Force. The envisioned goal of this task force is to support the research community and policy makers to keep abreast of relevant research and drug development in COVID-19. One approach to this is looking through the lens of clinical trials and tracking their design, progress and outcomes.

The Task Force has done further reviews and curated available information on clinical trials in India from March 2020 to December 2020. Additionally, data analysis of this curated data was done to help in designing further studies for COVID-19 management. The Task Force has released its reports via the website https://nclinnovations.org/covid19/resources/.^9^ This position paper compiles the data of interventional clinical trials registered on Indian CT registry, CTRI. An attempt has been made to analyse the data for comparing various trial parameters which gives us insightful conclusions and directions to move ahead in COVID 19 research.

## Materials and Methods

### Data Collection Methodology

Clinical trials related to COVID-19 were identified and data was collected from the Clinical trial registry of India (CTRI) database. CTRI, is operated by the National Institute of Medical Statistics, Indian Council of Medical Research (ICMR). Trials registered from March 2020 to December 2020 were considered for this purpose. Keywords used for identification comprised of COVID, SARS-CoV-2, Novel Coronavirus, Corona Virus or COVID-19. Since the objective of this data compilation was to support researchers in drug development process and assist policy makers in India it was decided to capture only interventional trials during this period and thus observational and Post marketing surveillance (PMS) trials were excluded.

The data retrieved from CTRI website included CTRI Number, Public Title, Type of Study (prophylaxis/ therapy), Type of Trial (observational/ Interventional), Recruitment Status, Completion status, Health Condition, Stage of the disease, Hospitalized (Y/N/Not mentioned), Inclusion Criteria, Exclusion criteria, Principal Investigator, Type of Sponsor, Sponsor, Intervention Name, Route of Administration, Drug, Details of Ayurveda/ Homeopathic Drug, Location, State, Study Design, Study design code, Start Date, End Date, Duration (months), Phase, Size, Dose, Primary Outcome, Time Point, Secondary Outcome and Time Point.

### Data analysis

The data was cleaned to remove any duplicate or erroneous entries before it was analysed using summary and descriptive statistics.

## Results

### General characteristics

Overall, during the study period 409 interventional clinical trials were registered for testing therapeutic benefits of drugs or plasma, vaccines, AYUSH products, Yoga and other products like nutraceuticals or Vitamins. It was found that 108 of the 409 registered trials were related to interventional trials on modern drugs, including repurposed drugs, convalescent plasma, monoclonal antibodies and vaccines with 83 trials focused on the repurposed drugs. Further, 67% of the trials were treatment related, 29% were prophylactic and 4% studied both the effects. Since our focus was clinical trials for modern drugs in India, we calculated the month-on-month percentage of trials on modern drugs as presented in Figure 1 below.

**Fig. 1:**
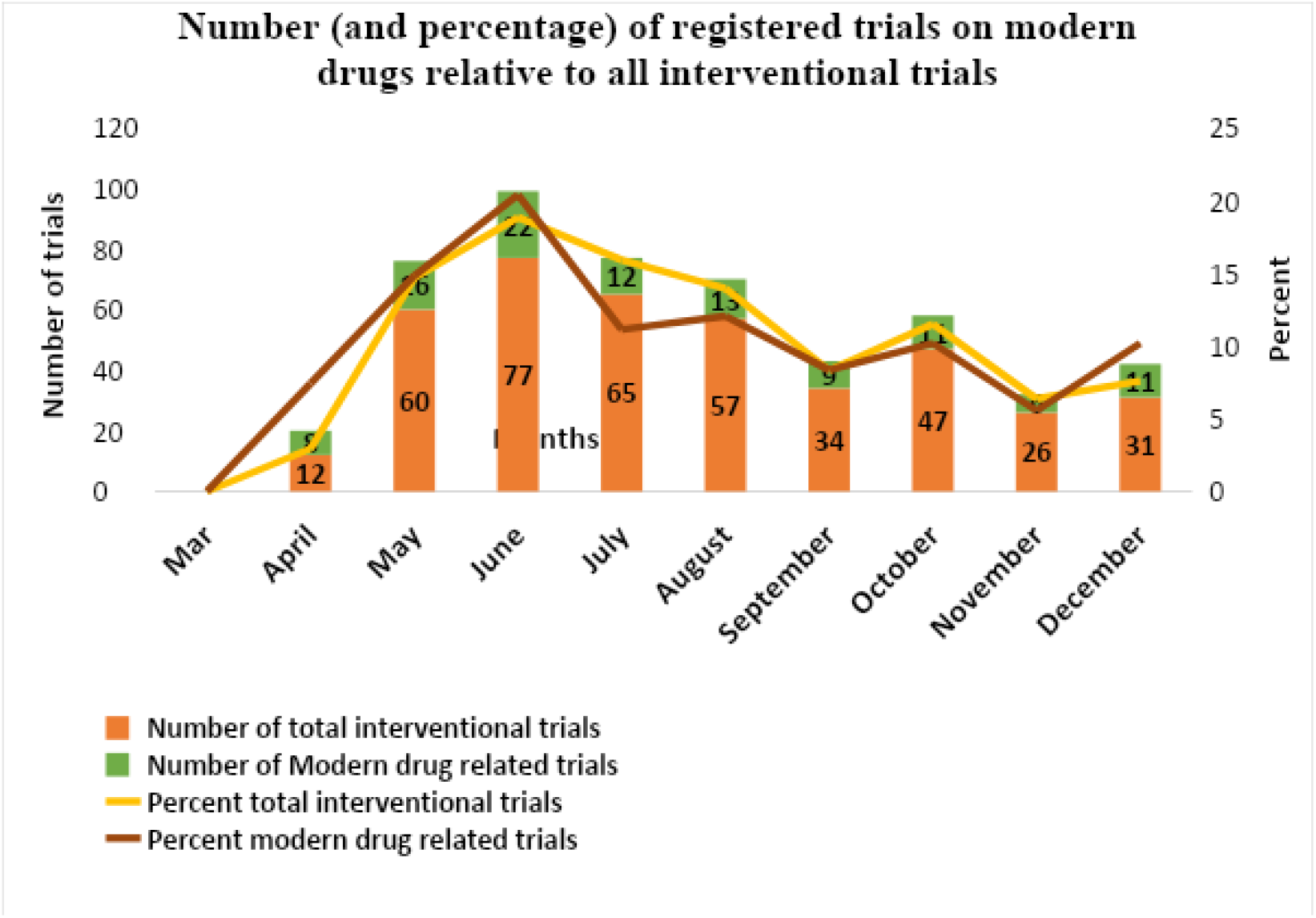
Trials on modern drugs relative to all interventional trials per month

**Fig. 2:**
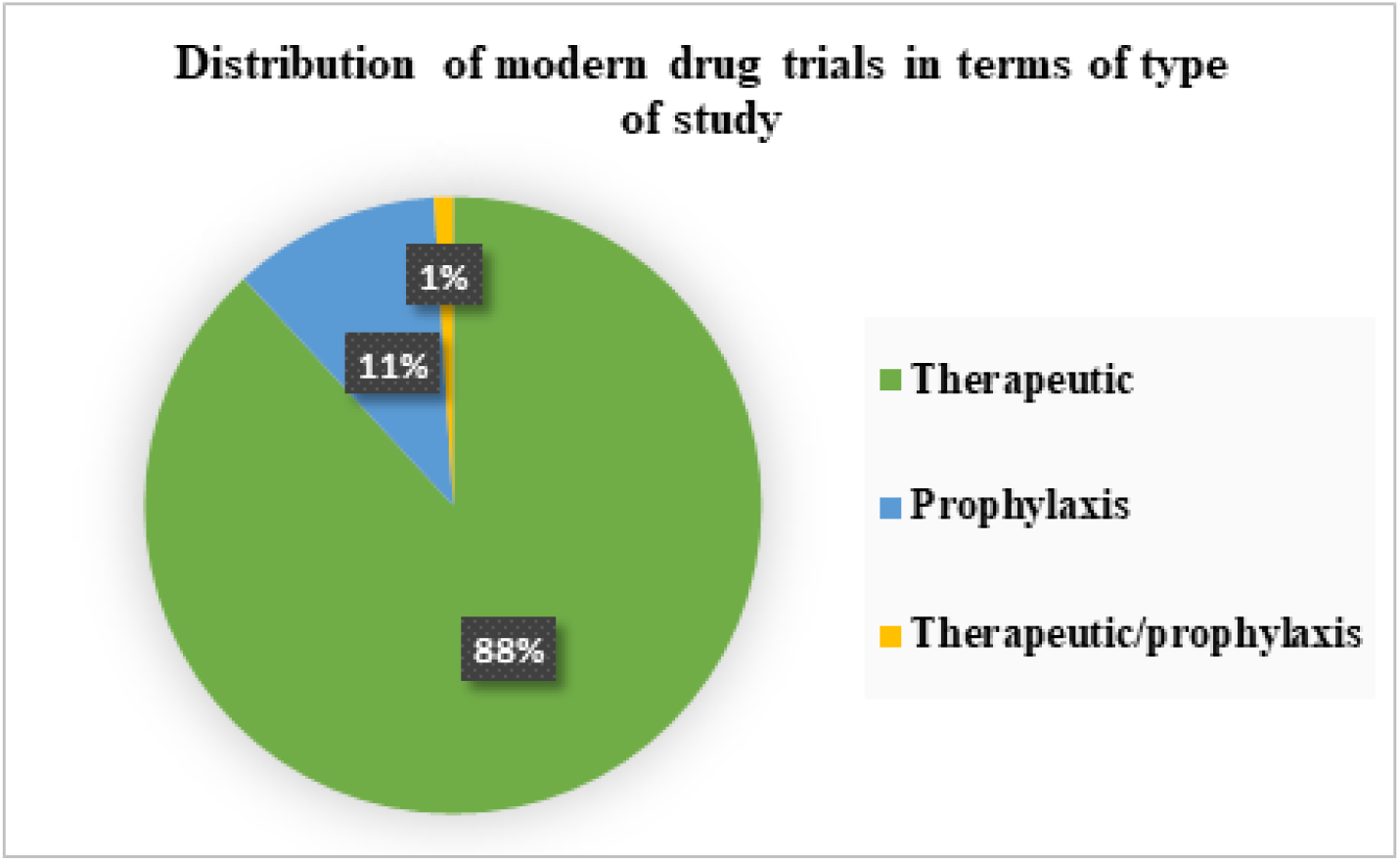
Distribution of modern drug trials in terms of type of study

It was also found that out of 108 registered trials on modern drugs,88% trials were therapeutic, 11% prophylactic and 1% with both the objectives (Fig.2).

We felt the need to understand if the pace of clinical research in India was matching the need for urgent promising interventions considering the growing number of cases and deaths each month. Fig. 3 compares the cumulative COVID-19 cases in India from March 2020 to Dec 2020 and number of clinical trials registered during this period. For data on cumulative number of cases/month we referred to the data available on https://www.statista.com/statistics/1104054/india-coronavirus-covid-19-daily-confirmed-recovered-death-cases/ accessed on 28^th^ January 2021^10^.

**Fig. 3:**
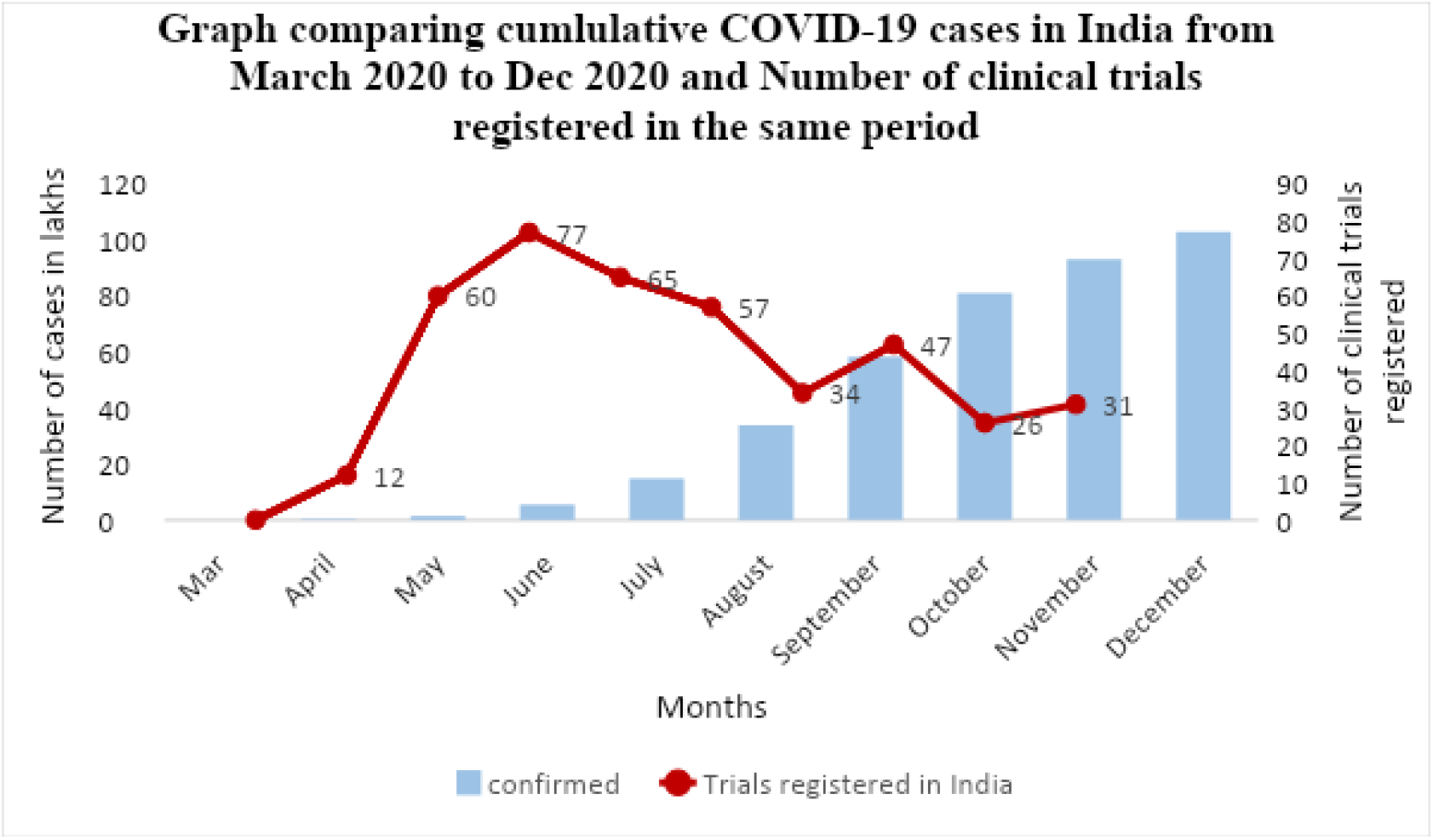
Graph comparing cumulative COVID-19 cases in India from March 2020 to Dec 2020 and Number of clinical trials registered in the same period

To achieve the objective of preparing India for a future pandemic, it is necessary to understand the study design pattern in the existing COVID-19 studies. Hence, we performed analysis of few study design related parameters.

As sample size plays a crucial role in interpreting the results of a trial, it was important to understand how the recruitment matched the trials registered.

Figure 4 clearly highlights that the number of trials registered increased from 60 in the month of May 2020 to 77 in the month of June 2020. However, proposed recruitment was seen to drop down in June 2020 compared to May 2020. Also, it is found that in the month of September 2020 when number of cases increased exponentially in comparison to August 2020 (Fig. 3) there is a drastic decrease in the number of trials registered (57 in August 2020 to 34 in September 2020) and also in the proposed recruitment (278197 in August 2020 to 9986 in September 2020) (Fig. 4)

**Fig. 4:**
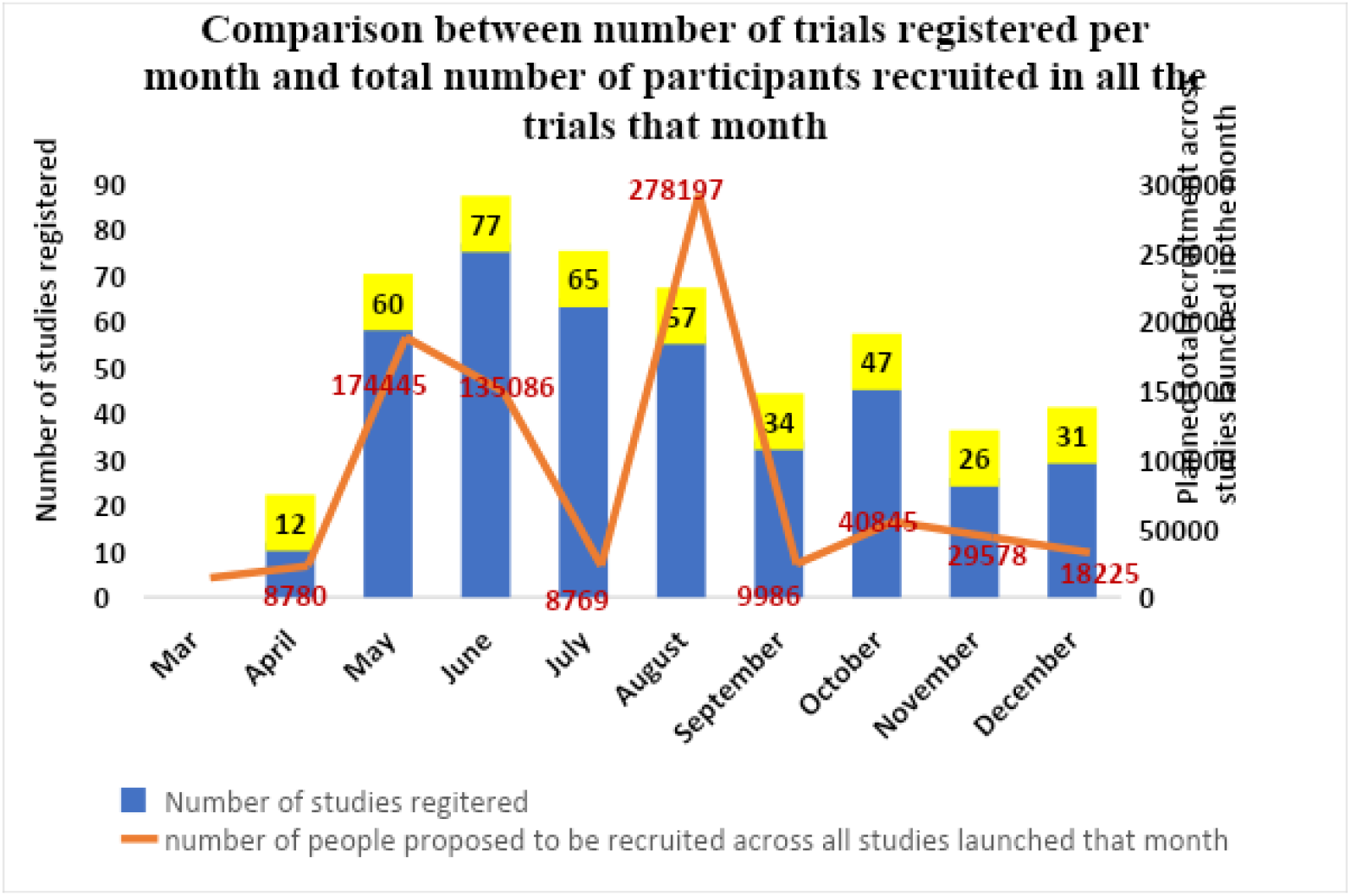
Number of registered trials and number of participants recruited in the trials for the period under study

### Analysis of Modern drug related trials

To track the pattern of clinical trial progress in the segment of modern drugs, we did a comprehensive and separate analysis of 108 trials. We analysed the available data for parameters like study design (Random/Non-random, sample size, phase of the study, duration of the study), recruitment status, primary sponsor and drug categories.

From the 108 trials studied, 92 trials were found to be randomized trials and 5 non randomized trials. There were 18 trials suggesting the use of placebo arm and 22 trials were active controlled trials. Data showed that for 79 trials recruitment status in India was “Not yet recruiting” and only 10 trials were “Open to recruitment”. Remaining 19 trials showed the status as “Not Applicable”. For any clinical trial, it is very important to have a consistent funding support. Analysis of drug related trials revealed that of 108, thirty-four trials were sponsored by Indian Pharmaceutical industry, 23 trials were self-sponsored and 20 trials were sponsored by the Research institute and hospital. Distribution amongst other primary sponsors is shown in table I.

**Table I:** Type of sponsors and Number of trials sponsored.

| Type of sponsor | Number |
| --- | --- |
| Private medical college | 3 |
| Private hospital/clinic | 5 |
| Other (state govt. health dept) | 6 |
| Govt. medical college | 12 |
| Govt. funding agency | 3 |
| Blank/Sponsor could not be identified | 2 |

**Note:** Other category sponsors include the State Govt. health dept, International Research Organisations, Indian Air Force, Charitable Health hospitals and Armed forces medical services

Achievement of the primary outcome in any clinical trial mainly depends on the study duration and planned sample size. Fig. 5 highlights 32 out of 108 trials were designed for 6 months followed by 17 trials for 3-month duration and 20 trials for 12-month duration.

**Fig. 5:**
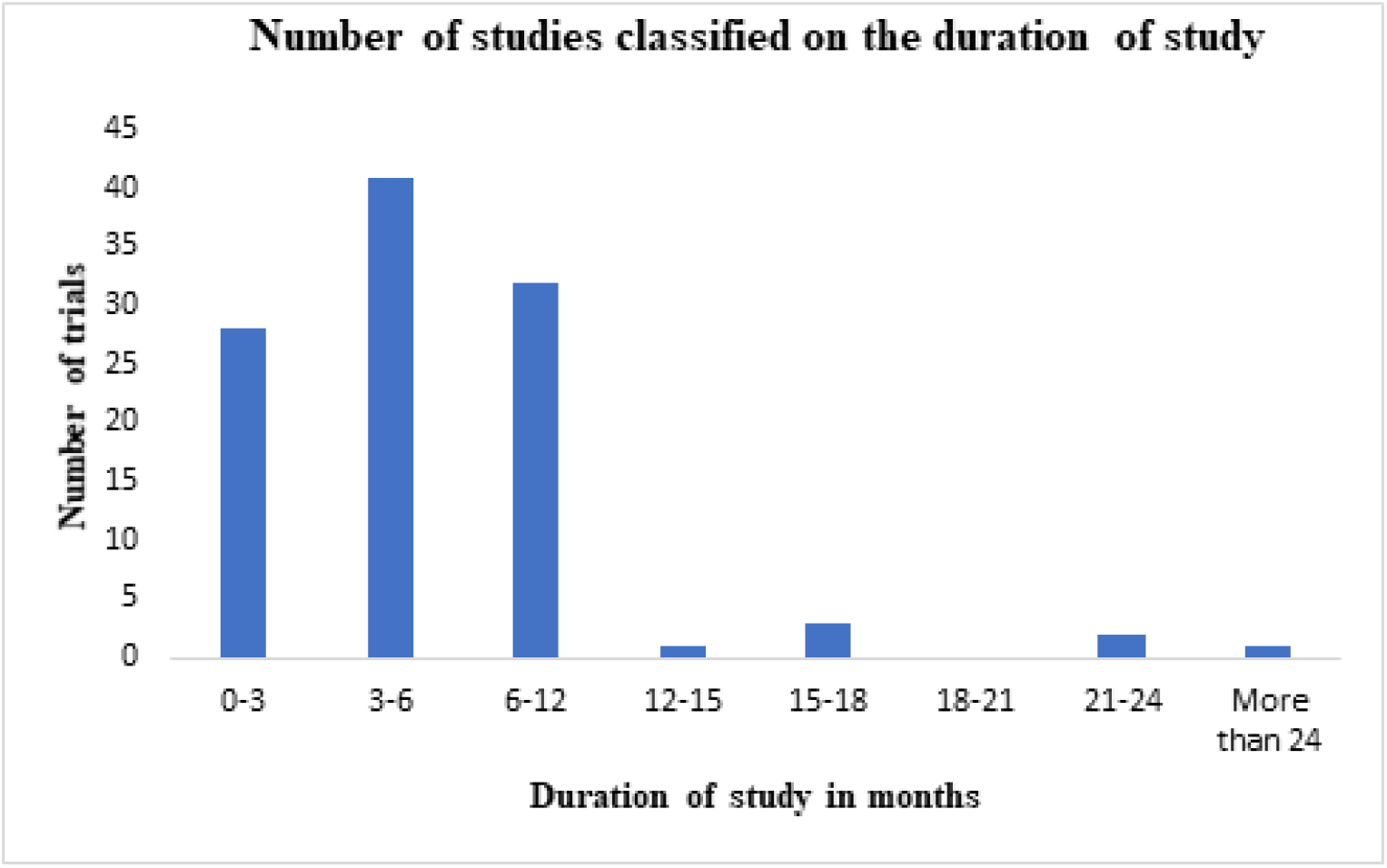
Number of studies by duration

Since sample size varies for each study, we decided to present this data in terms of the range of the planned size. Table II shows that 30 registered trials had a sample size range between 50-100 followed by 27 trials with size range 10-200 and 25 with 0-50 as size range.

**Table II:** Number of trials in defined Sample size range.

| Sample size range | Number |
| --- | --- |
| 0-50 | 25 |
| 50-100 | 30 |
| 100-200 | 27 |
| 200-300 | 8 |
| 300-400 | 4 |
| 400-500 | 4 |
| 500-600 | 2 |
| 600-700 | 1 |
| 700-800 | 0 |
| 800-900 | 1 |
| 900-1000 | 1 |
| > 1000 | 5 |

However, looking at study duration and sample size separately is of limited significance. We also looked at duration and sample size together. We found that 13 out of 32 trials with 6-month duration had a sample size range between 100-200, 9 out of 20 with 12-month duration had a sample size range between 100-200 and 11 out of 17 with 3-month duration had a sample size range between 0-50. As sample size has relevance with trial phase, we also studied distribution of size ranges in each of the trial phases (Fig 6 and Fig. 7). It shows that highest number (38 out of 108) trials were conducted as Phase 2 with 10 trials having sample size range 0-50, 11 with size range 50-100, 9 with size range 100-200, 3 with size range 200-300, 2 with size range 400-500, 1 with 500-600 and 1 with more than 1000.

**Fig 6:**
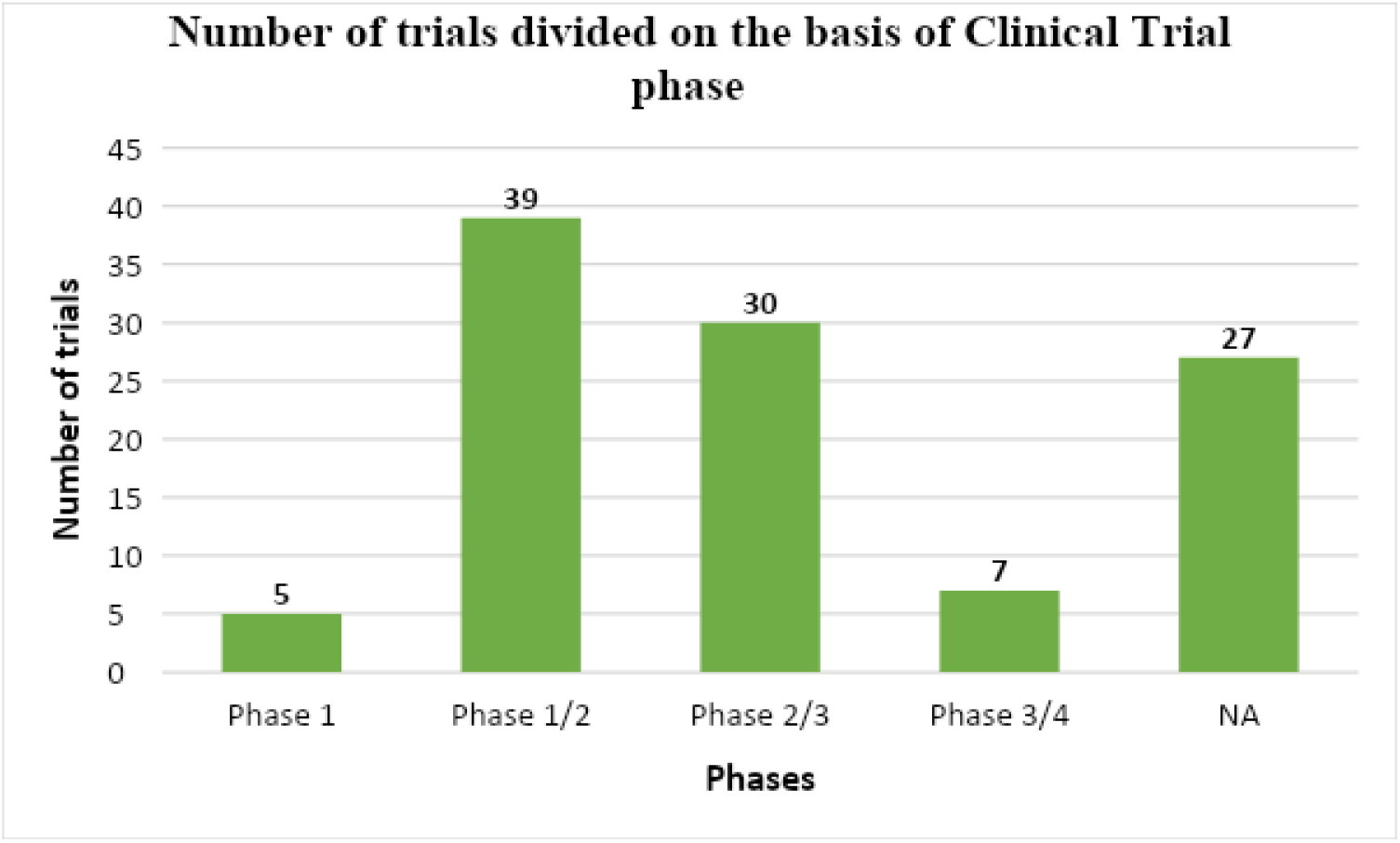
Number of trials by Clinical Trial phase

**Fig. 7:**
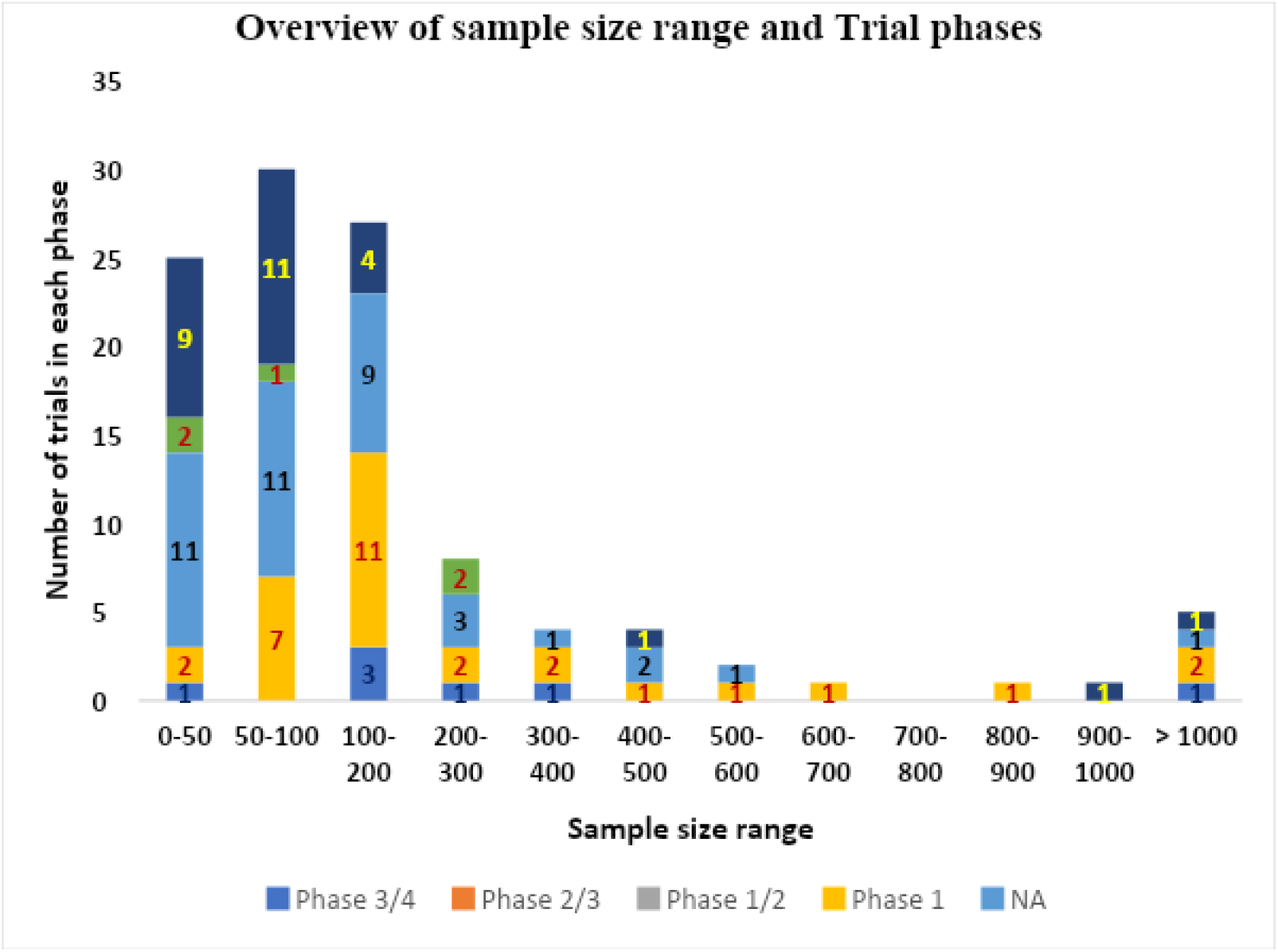
Overview of sample size range and trial phases

Lastly, we analysed the data on the basis of drug categories. Data showed that only 30% trials are registered for efficacy of anti-virals whereas remaining 70% focus on other treatments like monoclonal antibodies, vaccines, Vitamins, anti-inflammatory agents and steroids.

## Discussion

This study focuses on the interventional trials registered with CTRI between March 2020 to December 2020. The study characterized the scope, objectives and content of the current clinical research in the Indian context. In interventional trials for safety or efficacy the participants were patients or healthy human volunteers depending on the primary outcome. Our analysis offers a reason for both sanguinity and attention.

### General characteristics

From the total of 409 trials registered, approximately 26% trials (108) focused on modern drugs. This indicates that 74% of the trials in India focused on AYUSH products, Yoga, other products like nutraceuticals, Vitamins or other alternative forms of medicine. This seems to be an encouraging finding and the Ministry of AYUSH^11^ is making judicious efforts to inspire researchers to explore these products for COVID 19. However, the aggressive need of finding effective treatment in the current situation demands increased efforts to perform clinical research on modern drugs as well. Considering the time-consuming drug development process, the rational approach adopted worldwide is repurposing of drugs. Our observations reveal that only 83 from 108 trials on modern drugs studied the repurposed drugs while the remaining 25 focussed on plasma therapy or new monoclonal antibodies.

A positive finding from these trials when analysed on the basis of type of study shows efforts were focussed on both therapeutic candidates and prophylactic agents.

Optimal sample size is an indispensable factor of any research. In a clinical trial, a small sample size may fail to answer the study hypothesis or may not be able to distinguish important effects and associations^12^. Looking at the number of trials registered in the study period, we evaluated the month-wise proposed cumulative recruitment in total trials registered. It showed that though the number of trials registered in the month of June was 77 as against 60 in the month of May, proposed recruitment got down from 24.8% to 19.2%. In contrast in the month of August total proposed recruitment was 39.5% in comparison to July (1.2%) when actually trials registered in August were lower (57) than in the month of July (65). It is common to have a lag of around 2-3 months before the recruitment starts due to activities related to initiation of the trial. Hence it is not surprising that the proposed recruitment percentage in August is high, in line with the number of trials registered in May and June. In addition, there are also known reasons for the actual recruitment rates being low. A study by Nick G Cunniffe^13^ and his colleagues states that feasible recruitment rates, study design, exclusion criteria and proliferation of trials can limit the number and size of trials. Other reasons for declining recruitment usually seen in any trial could be a result of specific protocol issues, inconvenience, other unknown reasons, financial constraints or participation in other trials. This was put forth by Brintnall-Karabelas^14^ and his colleagues. We could not access data on actual versus planned recruitment and hence could not corroborate these observations. However, with all these issues known to hamper recruitment in trials, and given that a large majority of the registered trials are not yet recruiting, it is desirable to consider fewer but more suitably designed trials to maximise the success rate in this need of emergency. Welcome-RECOVERY trial^15^ and WHO-Solidarity trials^16^ could be the guiding trials to understand the key aspects of trial design while addressing such emergencies.

### Modern drug related trials

We filtered modern drug related trials from the total set of interventional trials to perform further analysis. This was necessary to attain objectives of the study initiated by the Task force for repurposing of drugs for COVID 19. Our main focus was understanding various aspects of study design.

Observations on 108 registered trials on modern drugs suggest that 85% (92) trials were designed as randomized trials and 4% (5) as non-randomized trials while remaining fall under the category of case studies or single-arm trials. This positive finding matches with the similar findings by Mehta and his colleagues^8^. Undoubtedly non-randomized or single arm trials or case studies provide an early indication of the probability of drug success, while randomised study designs provide better quality evidence to arrive at effective and safe treatments. In context of COVID 19 trials in India, however, it is seen that standard of care is followed in both treatment as well as control arm. As a result, the ethical issue of placebo-controlled trials is not relevant. However, the major finding of concern is that only 10 out of the 108 modern drug related trials were open for recruitment, while 79 showed status as not yet recruiting (and 19 showed status as Not Applicable). There is an urgent need to investigate why such a high number of trials were not initiated. One reason which we could relate to this was our next observation regarding primary sponsors. We found that Indian pharmaceutical industries, research institutes and hospitals were the top primary sponsors whereas a limited number of trials were sponsored by Government-funded agencies. Similarly, very few studies had identified secondary sponsors. This is in contrast with the two large trials Welcome-RECOVERY and Solidarity^15,16^ which were sponsored by NIHR and WHO respectively along with industry and academic partners. The success of RECOVERY and Solidarity trials as seen through their interim results reinforces the value of designing a single, large, statistically sound, multi-centre trial covering multiple interventions in parallel. This will allow pooling of funds from various agencies to initiate one promising trial.

We also tried to understand if the study durations and sample sizes of various trials across phases were suitable to achieve the primary outcomes.

As mentioned in an article by Pourhoseingholi^17^ in a phase I trial that involves drug safety on health, human volunteers total of around 20-80 patients may be considered. Phase II trials that investigate the drug efficacy, would require 100-200 patients.

The sample size calculation should be based on the primary outcome^18^. Data exhibited 30 trials to be Phase 2/3 trials. From these 11 trials were shown to have sufficient sample size ranging between 100-200 patients, 7 trials with sample size range 200-900, 2 trials with more than 1000 patients and 9 trials with less than 100 patients. Overall sample size selection was found to be satisfactory considering the phase of a trial however we could not establish any relation between study duration considered and sample size selected.

Lastly, we would like to mention that out of the 108 trials studied for modern drugs, mere 30% trials focused on evaluating anti-viral agents. Antiviral treatment will remain an extremely important component of the treatment of COVID 19 in spite of the successful vaccines coming up and large vaccination drives that are being rolled out in various parts of the world. The need for promising antivirals acting through multiple mechanism of action becomes crucial especially during the early stage of the disease, for resistant strains of the virus and for the population which cannot be vaccinated. As mentioned aptly in an article published by Zhaori *et. al*^19^ “there is a need to consider and evaluate which drugs are truly antiviral agents, to reach a clear conclusion about which drug(s) are effective and safe in treatment and prevention of this pandemic disease and worthy of recommending for clinical application”. Though the context of Zhaori’s manuscript points to the paediatric segment, it is applicable to the general population too.

## Conclusions and Way forward

Through this study we have tried to document the existing or ongoing clinical trials in India on COVID-19 between March 2020 to December 2020. Large number of interventional trials covering a large number of prospective subjects were carried out in India. Majority of it were for traditional medicine (74%) and around 26% were for modern drugs. An attempt has been made to present the consolidated analysis of the interventional trials for modern drugs w.r.t their design, sample size, recruitment status, primary sponsors etc. This analysis will assist researchers, decision makers and clinicians to comprehend the clinical research situation in India. We strongly feel that the Government, Academic institutions, Research institutions and Indian Pharmaceutical industries should not be working in silos but should indulge into continuous dialogue to fight this common cause. This way India may also contribute towards the effort to identify drugs to fight against the COVID 19 at the global level. The study is also useful for the global scientific community as a potential source of information for international collaborations. India being a major hub for clinical research, a joint effort by the relevant stakeholders can help overcome the current lacunae in the clinical trials.

## Data Availability

Clinical Trials for Covid19 in India- data till 30 Nov 2020

https://docs.google.com/spreadsheets/d/188Bkbb16MaRg8VGvsFH9J3QQ_-SxyGgP9nUIFIXANog/edit?ts=5f5370af#gid=159465981

## Acknowledgements

The authors are grateful to the Department of Scientific & Industrial Research (DSIR) for funding the work under A2K+ Scheme of DSIR. The authors are also thankful to the advisory group of the Task Force for Repurposing of Drugs (TFORD) for COVID-19, under the aegis of the Science and Technology core group.

